# Fluctuations in serum adrenocorticotropic hormone concentration may predict the onset of immune checkpoint inhibitor-related hypophysitis

**DOI:** 10.1101/2023.12.05.23299427

**Authors:** Hironori Bando, Masaaki Yamamoto, Shin Urai, Yuma Motomura, Yuriko Sasaki, Yuka Ohmachi, Masaki Kobatake, Yasutaka Tsujimoto, Yuka Oi-Yo, Masaki Suzuki, Naoki Yamamoto, Michiko Takahashi, Hidenori Fukuoka, Genzo Iguchi, Wataru Ogawa

**Author notes:** **Correspondence:** Hironori Bando M.D, Ph.D., Division of Diabetes and Endocrinology, Department of Internal Medicine, Kobe University Hospital, 7-5-1, Kusunoki, Chuo, Kobe, 650-0017, Japan.

## Abstract

**Background:** Immune checkpoint inhibitor (ICI)-related hypophysitis (RH) is a common immune-related adverse event. The early detection of ICI-RH prevents life-threatening adrenal insufficiency. However, good predictors of secondary adrenal insufficiency in ICI-RH have not yet been reported. We hypothesized that fluctuations in serum adrenocorticotropic hormone (ACTH) and cortisol levels occur similarly to those in thyroid-stimulating hormone and thyroid hormones (thyroxine and triiodothyronine) in ICI-related thyroiditis. Here, we sought to test this hypothesis.

**Methods:** Patients who used ICI and had a history of measurement of serum ACTH and cortisol concentrations were retrieved from electronic medical records, and those with a history of glucocorticoid use were excluded from the analysis. We evaluated fluctuations in serum ACTH and cortisol concentrations and the development of ICI-RH. For patients with ICI-RH, data at three points (before ICI administration [pre], maximum ACTH concentration [peak], and onset of ICI-RH) were analyzed to evaluate hormone fluctuations.

**Results:** A total of 202 patients were retrieved from the medical record. Forty-three patients were diagnosed with ICI-RH. Twenty-six out of 43 patients had sufficient data to evaluate fluctuations in serum ACTH and cortisol concentrations and no history of glucocorticoid use. ACTH concentrations changed from 37.4 [29.9–48.3] (pre) to 64.4 [46.5–106.2] pg/mL (peak) (1.72–fold increase, *p* = 0.0026) in the patients with ICI-RH before the onset. There were no differences in cortisol concentrations between the pre and peak values in patients with ICI-RH. We also evaluated the fluctuations in serum ACTH and cortisol levels in patients who did not receive ICI-RH (62 cases). However, elevation of serum ACTH levels was not observed in patients without ICI-RH, suggesting that transient elevation of serum ACTH is a unique phenomenon in patients with ICI-RH.

**Conclusions:** Serum ACTH levels were transiently elevated in some patients with ICI-RH before the onset of secondary adrenal insufficiency. Monitoring the ACTH levels and their fluctuations can help predict the onset of ICI-RH.

## Background

Immune checkpoint inhibitors (ICIs) are widely used to treat various malignant neoplasms. Programmed cell death protein 1 (PD-1), programmed death ligand 1 (PD-L1), and cytotoxic T-lymphocyte-associated protein 4 (CTLA-4) are the ICI targets. However, ICI treatment can lead to immune-related adverse events (irAEs), including autoimmune endocrinopathies. Pituitary insufficiency due to ICI-related hypophysitis (ICI-RH) is a common irAE. The development of pituitary insufficiency in patients receiving ICIs is associated with better overall survival in several malignancies(1). However, the appropriate diagnosis, early detection, and treatment of hypopituitarism are essential. Adrenal insufficiency due to hypopituitarism can be lethal if not properly managed. Life-threatening adrenal insufficiency is characterized by hypotension, dehydration, hyponatremia, and hyperkalemia, which mimics sepsis, brain metastasis, and cachexia(2). The frequency and timing of onset vary with ICI class. The frequency of ICI-RH in anti-CTLA-4 antibody (Ab)-based regimens is 5–10% with a median onset of 9–12 weeks, and that in anti-programmed cell death-1 (PD-1) and programmed cell death ligand 1 (PD-L1)-based regimens is <1% with a median onset of 26 weeks(3). However, the timing of disease onset varies considerably among cases, hindering the prediction of disease onset.

The clinical features of anti-CTLA-4 and anti-PD-1/anti-PD-L1 Ab-RH differ. Anti-CTLA-4 Ab-RH causes panhypopituitarism, including secondary hypothyroidism, hypogonadotropic hypogonadism, and secondary adrenal insufficiency(4). In contrast to anti-CTLA-4 Ab-RH, anti-PD-1/PD-L1 Ab-RH predominantly causes an isolated adrenocorticotropic hormone (ACTH) deficiency(5). These different patterns may result from different underlying mechanisms(6). However, secondary adrenal insufficiency accounts for 83% of ICI-RH cases(7). Therefore, predictive markers for the onset of secondary adrenal insufficiency are essential.

Tahir et al. reported autoantibodies against guanine nucleotide-binding protein G(olf) subunit alpha (GNAL) and integral membrane protein 2 B (ITM2B) as biomarkers for ICI-RH(8). However, these biomarkers are still under investigation and cannot be measured routinely. Therefore, markers that can predict the onset of hypopituitarism under regular measures are necessary. Several studies have reported an elevation of the eosinophil fraction and count during the last visit and at the onset of hypopituitarism(9, 10). These studies raised a crucial point; however, this report also showed that the cortisol concentration had already mildly decreased at the last visit, which may be seen as a sign that ICI-RH is already under development. Eosinophilia is a well-known marker of already developed adrenal insufficiency(11).

Thyroiditis is one of the most common irAEs. ICI-related thyroiditis typically occurs biphasically: thyrotoxicosis, followed by a subsequent persistent hypothyroidism(12). In the initial thyrotoxicosis phase, free thyroxine and triiodothyronine levels are elevated while those of thyroid-stimulating hormone (TSH) are suppressed. In the hypothyroidism phase, the pattern is reversed. The underlying mechanism of ICI-related thyroid dysfunction remains unknown. We speculated that, if the pituitary gland is damaged in ICI-RH by a mechanism similar to that of thyroiditis, serum ACTH and cortisol levels, which are indicators of the hypothalamic pituitary adrenal (HPA) axis in routine practice, may fluctuate during the process. If so, they could potentially be used as predictive factors for the onset of secondary adrenal insufficiency in ICI-RH. To explore this possibility, we collected data on serum ACTH and cortisol levels in patients using ICIs and analyzed fluctuations in hormone concentrations.

## Methods

### Patients

To avoid arbitrarily selecting and detecting patients with ICI-RH from the electronic medical records of Kobe University Hospital, we extracted patient data using the following criteria: 1. use of ICI(s) at least once and 2. measured plasma ACTH and cortisol levels at least twice. We thought that if ACTH and cortisol concentrations were measured at least twice, we could identify the patients who developed ICI-RH. The data extraction period for starting the ICI treatment was from April 2014 to March 2022. The follow-up period was until October 2023. The reason for the follow-up duration was that 18 months had passed since the initiation of ICI treatment, and most ICI-RHs are developed by that time(13). After retrieval, we checked the patients’ medical records and diagnoses of ICI-RH. Some chemotherapy protocols that combine ICI and cytotoxicity require exogenous glucocorticoids as adjuvants to relieve side effects. Exogenous glucocorticoids suppress the HPA axis, challenging the evaluation of the ACTH and cortisol concentrations to correctly diagnose hypopituitarism. Therefore, patients with a history of glucocorticoid use were excluded from our analysis. We also excluded patients who regularly used a pharmacological dose of glucocorticoids for autoimmune or allergic diseases from the ACTH and cortisol concentration analyses for the same reason. Thus, only cases of ICI-related hypopituitarism in which ACTH and cortisol fluctuations could truly be evaluated were selected. Cases in which glucocorticoids made it challenging to assess ACTH and cortisol levels were excluded.

### Data collection and analysis

After selecting patients who were truly diagnosed with ICI-RH, we analyzed the morning ACTH and cortisol concentrations at three points during the clinical course: 1. just before the initiation of ICI therapy (pre), 2. the timing that showed the maximum ACTH concentration after initiation before the onset of ICI-RH (peak), and 3. the onset of ICI-RH. Therefore, we excluded data that lacked the abovementioned three points for ACTH and cortisol levels. Data on the Na, K, and eosinophil fractions at the abovementioned points were also collected. We also analyzed the data of patients who did not develop ICI-RH (non-ICI-RH). ACTH and cortisol concentrations were analyzed at two different time points: 1. just before the initiation of ICI (pre), and 2. the timing showing the maximum ACTH concentration after initiation of ICI (peak).

### Statistical analysis

Statistical analyses were performed using JMP Statistical Database Software version 12·2·0 (SAS Institute, Cary, NC). The Mann-Whitney U test and Wilcoxon signed-rank test were performed as appropriate. Statistical significance was set at *p* < 0.05.

## Results

Using the criteria mentioned above, the electronic medical records of 202 patients who used ICIs at least once and whose serum ACTH and cortisol concentrations were measured at least twice were obtained. Among them, 43 patients were diagnosed with ICI-RH. All patients in our cohort with ICI-RH had only reduced ACTH and cortisol levels, while the levels of other pituitary hormones were unaffected, resulting in the diagnosis of isolated ACTH deficiency. Isolated ACTH deficiency was diagnosed as previously described(14). Five patients were excluded owing to a history of glucocorticoid use as an adjuvant. One patient who used pharmacological glucocorticoids to treat allergic diseases was also excluded from the analysis. Eleven patients lacked ACTH or cortisol data just before the initiation of ICI and/or the peak ACTH concentration after initiation, before the onset of ICI-RH. Similarly, for non-ICI-RH cases, we excluded patients who were inappropriate for analysis because of a history of steroid use or lack of data. Finally, we analyzed the fluctuations in ACTH and cortisol concentrations in 26 and 62 patients with ICI-RH and non-ICI-RH, respectively (Table 1).

**Table 1.** Patient characteristics.

|  |  | **ICI-RH cases** |  | **Non-ICI-RH cases** |
| --- | --- | --- | --- | --- |
| **number of patients** |  | 26 |  | 62 |
| **age of ICI therapy initiation** |  | 68 [62–71] |  | 70 [65–74] |
| **sex (male and female)** |  | 19 and 7 |  | 49 and 13 |
| **primary organ of the tumors\*** | kidney | 12 | kidney | 14 |
|  | lung | 6 | lung | 15 |
|  | pharynx | 4 | pharynx | 13 |
|  | skin | 2 | skin | 5 |
|  | esophagus | 2 | esophagus | 6 |
|  | stomach | 2 | stomach | 4 |
|  | uterus | 1 |  |  |
|  | unknown primary<br>origin | 1 | unknown primary<br>origin | 3 |
|  |  |  | urinary duct | 3 |
|  |  |  | bladder | 2 |
|  |  |  | parotid gland | 1 |
|  |  |  | liver | 1 |
| **ICI-RH**<br>**complications**<br>**(n, %)** |  | 5, 19.2% |  | 13, 20.1% |
| **ICI type<sup>#</sup>** | nivolumab | 15 |  | 32 |
|  | pembrolizumab | 5 |  | 16 |
|  | ipilimumab +<br>nivolumab | 5 |  | 6 |
|  | atezolizumab | 1 |  | 5 |
|  | durvalumab | 0 |  | 4 |

ICI: immune checkpoint inhibitor; RH: related hypophysitis
\*: includes multiple kinds of tumors

During the clinical course, fluctuations in the ACTH and cortisol concentrations were observed (Table 2, Figure 1). ACTH concentrations changed from 37.4 [29.9–48.3] (pre) to 64.4 [46.5–106.2] pg/mL (peak) (Figure 1A). ACTH concentrations were significantly elevated before ICI-RH onset after initiating ICI therapy (1.72-fold increase, *p* = 0.0026). The duration from ICI initiation to peak and from peak to ICI-RH onset was 49 [28–127] and 70 [39–186] days, respectively. There were no correlations between the pre and peak ACTH concentrations. Cortisol concentrations were statically not elevated at the peak of ACTH concentration (pre and peak concentrations were 11.8 [10.4–15.2] (pre) and 14.5 [11.5–16.5] (peak) μg/dL, respectively; *p =* 0.1742) (Figure 1B). The change in the ACTH/cortisol ratio was statistically significant (pre-and peak ratios were 3.30 [2.17–4.11] (pre) and 4.60 [3.33–7.01] (peak), respectively; *p =* 0.0070) (Figure 1C). We analyzed the eosinophil fraction at peak ACTH levels. We collected data on the eosinophil fraction from 22 of 26 patients. The eosinophil fraction at the time of peak ACTH was 2.8% [1.4–5.9%] (reference range: <7%) in this group. Only three patients showed an elevated eosinophil fraction (12.7, 13.0, and 25.0%).

**Figure 1.**
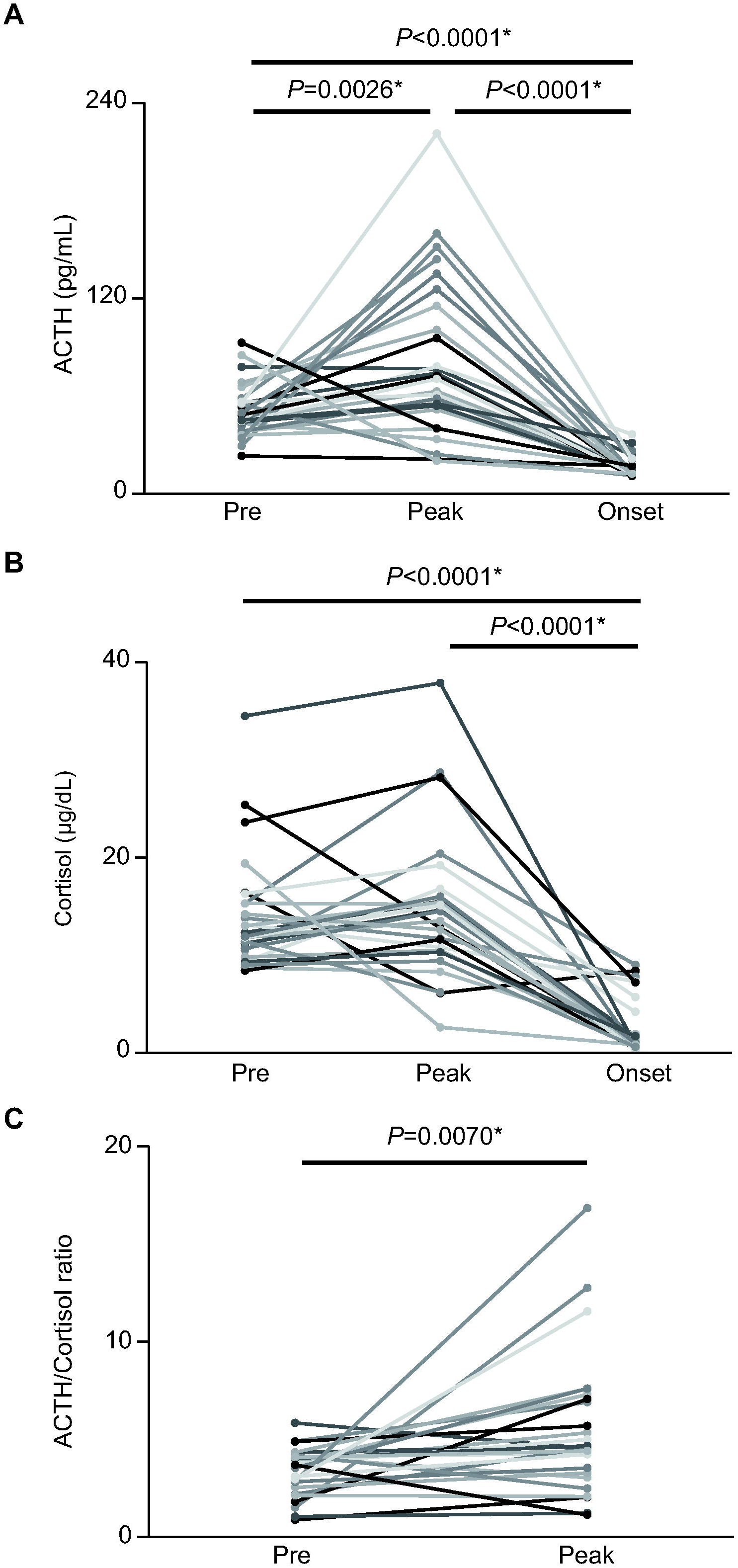
Fluctuations in ACTH and cortisol concentration and ACTH/cortisol ratio in patients with ICI-related hypophysitis (ICI-RH). (A) ACTH concentration, (B) cortisol concentration, and (C) ACTH/cortisol ratio.

**Table 2.**
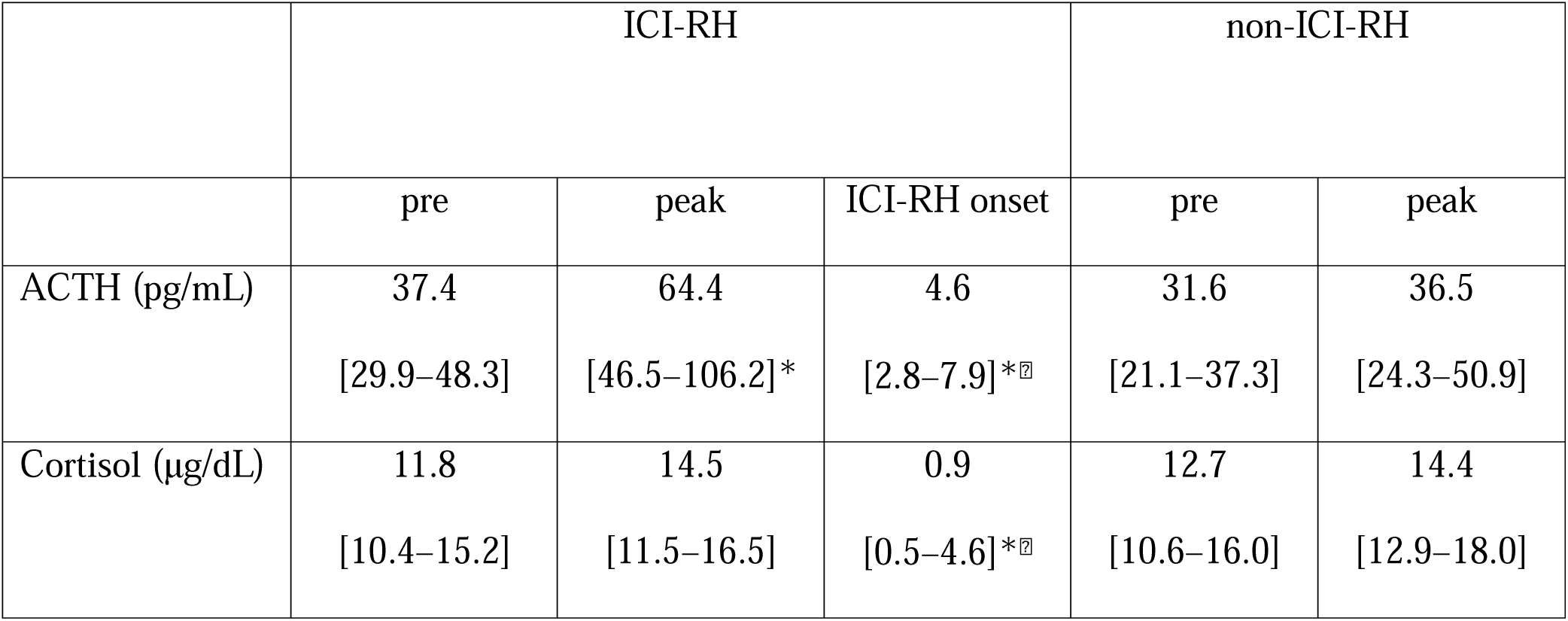
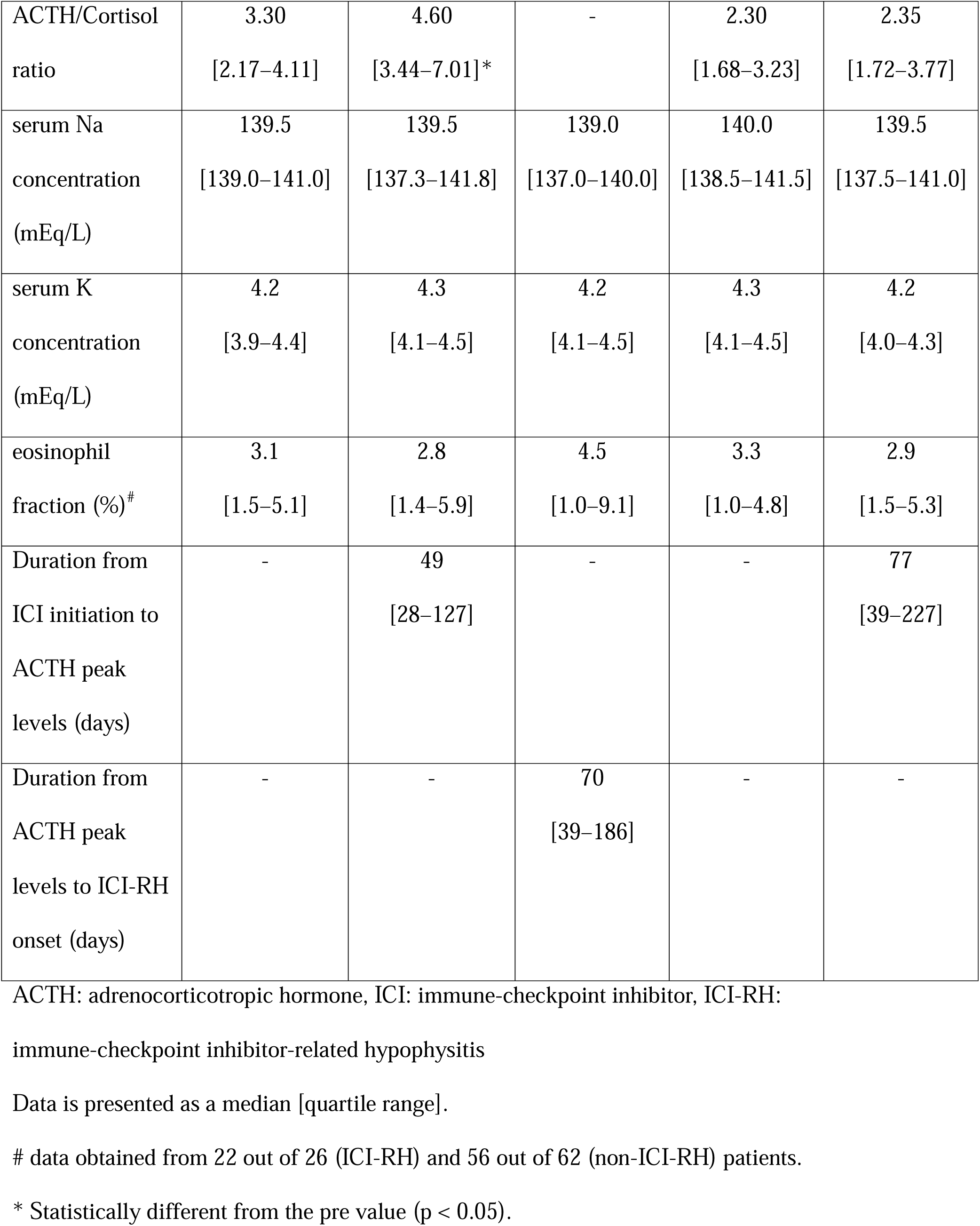
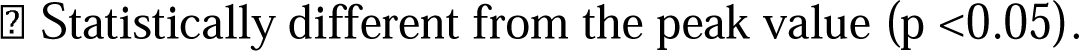
Fluctuations in ACTH and cortisol levels in patients with or without ICI-RH.

|  | ICI-RH |  |  | non-ICI-RH |  |
| --- | --- | --- | --- | --- | --- |
|  | pre | peak | ICI-RH onset | pre | peak |
| ACTH (pg/mL) | 37.4<br>[29.9–48.3] | 64.4<br>[46.5–106.2]* | 4.6<br>[2.8–7.9]*<sup>‡</sup> | 31.6<br>[21.1–37.3] | 36.5<br>[24.3–50.9] |
| Cortisol ($\mu\text{g/dL}$) | 11.8<br>[10.4–15.2] | 14.5<br>[11.5–16.5] | 0.9<br>[0.5–4.6]*<sup>‡</sup> | 12.7<br>[10.6–16.0] | 14.4<br>[12.9–18.0] |
| ACTH/Cortisol ratio | 3.30<br>[2.17–4.11] | 4.60<br>[3.44–7.01]* | - | 2.30<br>[1.68–3.23] | 2.35<br>[1.72–3.77] |
| serum Na concentration (mEq/L) | 139.5<br>[139.0–141.0] | 139.5<br>[137.3–141.8] | 139.0<br>[137.0–140.0] | 140.0<br>[138.5–141.5] | 139.5<br>[137.5–141.0] |
| serum K concentration (mEq/L) | 4.2<br>[3.9–4.4] | 4.3<br>[4.1–4.5] | 4.2<br>[4.1–4.5] | 4.3<br>[4.1–4.5] | 4.2<br>[4.0–4.3] |
| eosinophil fraction (%)<sup>#</sup> | 3.1<br>[1.5–5.1] | 2.8<br>[1.4–5.9] | 4.5<br>[1.0–9.1] | 3.3<br>[1.0–4.8] | 2.9<br>[1.5–5.3] |
| Duration from ICI initiation to ACTH peak levels (days) | - | 49<br>[28–127] | - | - | 77<br>[39–227] |
| Duration from ACTH peak levels to ICI-RH onset (days) | - | - | 70<br>[39–186] | - | - |

ACTH: adrenocorticotrophic hormone, ICI: immune-checkpoint inhibitor, ICI-RH:
immune-checkpoint inhibitor-related hypophysitis
Data is presented as a median [quartile range].
<sup>#</sup> data obtained from 22 out of 26 (ICI-RH) and 56 out of 62 (non-ICI-RH) patients.
\* Statistically different from the pre value ( $p < 0.05$ ).
□ Statistically different from the peak value ( $p < 0.05$ ).

Considering that the mechanism of pituitary dysfunction may differ between the ACTH-elevated and ACTH-non-elevated groups, we analyzed the clinical data of the groups. Since the median elevation of ACTH was a 1.72-fold increase, we divided the patients into two groups for analysis: the ACTH-increased group at the peak (>1.72) and the non-increased group (<1.72). However, there were no noticeable clinical differences (age; sex; type of malignancy; frequency of accompanying thyroiditis; concentrations of ACTH, cortisol, sodium (Na), and potassium (K); and eosinophil fraction at the onset of ICI-RH) (data not shown). All three patients with elevated eosinophil fractions at peak ACTH levels were included in the non-ACTH-elevated group.

Next, differences in fluctuations in ACTH and cortisol levels were evaluated in the non-ICI-RH group (Table 2). We collected data on ACTH and cortisol levels at two points: before the initiation of ICI therapy and at the time of highest ACTH concentration during the clinical course. Although no clear ACTH peak existed in the non-ICI-RH cases, the maximum ACTH concentrations measured during follow-up were used for analysis, as in the ICU-RH cases (duration from the initiation of ICI treatment to the peak was 77 [39–227] days). However, there was no elevation in ACTH, cortisol, or ACTH/cortisol ratio in patients without ICI-RH. We also evaluated the differences in the peak concentrations of ACTH and cortisol, and the ACTH/cortisol ratio between the ICI-RH and non-ICI-RH groups (Figure 2). The two groups had no noticeable differences in clinical backgrounds or observation periods. The peak ACTH concentration and ACTH/cortisol ratio, but not the cortisol concentration, of the ICI-RH group were statistically higher than those of the non-ICI-RH group, indicating that fluctuations in ACTH levels are specific to the ICI-RH group.

**Figure 2.**
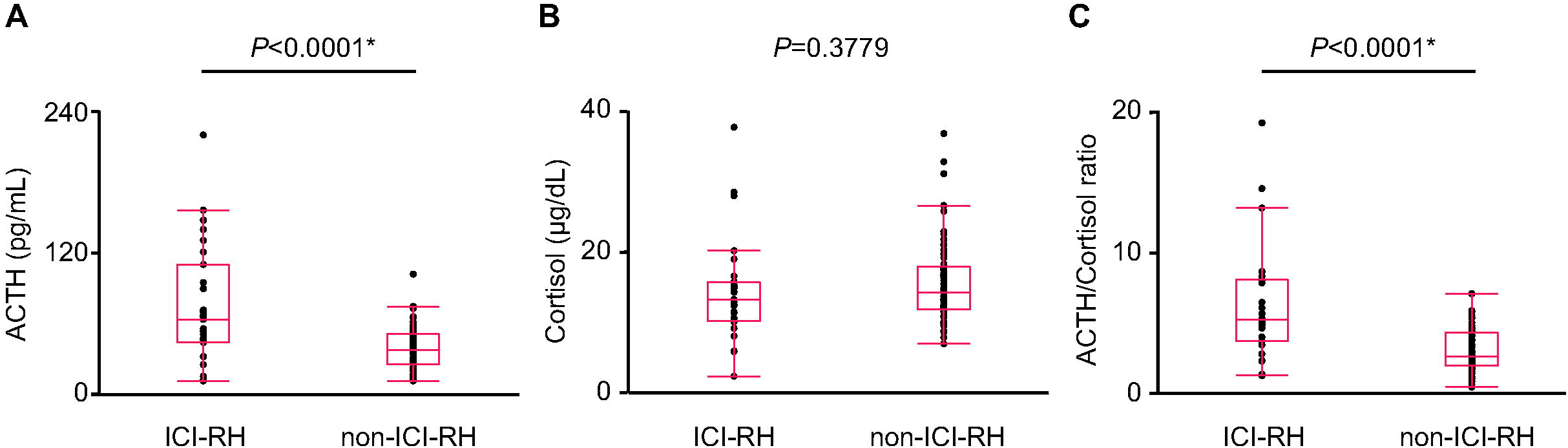
Differences in peak concentrations of ACTH and cortisol, and ACTH/cortisol ratio between ICI-RH and non-IC-RH groups during the clinical course. (A) ACTH concentration, (B) cortisol concentration, and (C) ACTH/cortisol ratio.

## Discussion

If the pituitary gland is damaged in ICI-RH by a mechanism similar to that of thyroiditis, serum ACTH and cortisol levels should fluctuate during the process and could be used as predictive factors for the onset of secondary adrenal insufficiency in ICI-RH. To explore this possibility, we collected data on serum ACTH and cortisol levels in patients using ICIs and analyzed fluctuations in hormone concentrations. ACTH levels were transiently elevated in patients who developed ICI-RH, and the elevation occurred prior to the onset of ICI-RH. These findings suggest that transiently elevated ACTH levels may be predictive of ICI-RH onset.

Glucocorticoid use suppresses the HPA axis, and ACTH levels are transiently elevated during the recovery of the HPA axis after suspension of chronic corticosteroid therapy(15). Since glucocorticoid use is common among patients receiving ICIs, only analyses omitting these cases can accurately examine the impact of ICIs on ACTH and cortisol levels as well as ICI-RH. Here, we analyzed patients in whom the effects of glucocorticoid use could be excluded. Hence, although the sample size was small, we could accurately evaluate the effect of ICI on ICI-RH onset and the concentrations of ACTH and cortisol. To our knowledge, this study is the first to show that a transient elevation in ACTH concentration occurs in some patients with ICI-RH. However, future studies are needed to determine the extent to which the transient elevation of ACTH levels is predictive of ICI-RH onset.

ACTH level elevation could occur prior to ICI-RH onset. There were no differences in the clinical characteristics between the ACTH-elevated and non-elevated groups. However, these two groups may have entirely different pathogenic mechanisms. We cannot rule out the possibility that the duration of elevated ACTH levels was extremely short and could not have been measured at the time of blood collection. Na and K levels did not change at the time of peak ACTH levels. Moreover, Na and K levels at the peak were similar between the ACTH-increased and non-increased groups. The significance of ACTH level elevation needs to be evaluated in future studies.

Three patients with elevated eosinophil fractions did not have elevated ACTH levels. In these cases, ACTH secretion may have been impaired when ACTH and cortisol levels were measured at the peak ACTH levels. After a transient rise in ACTH, ICI-RH onset occurs at a median of 70 days, but the time to onset varies widely. When this phenomenon is observed, attention to adrenal insufficiency symptoms may prevent adrenal crisis.

There are several possible explanations for ACTH level elevation without cortisol level elevation. One possibility is that a bioinactive form of ACTH may be abundant. ACTH is produced from proopiomelanocortin (POMC) and has several isoforms. ACTH(1-24) has steroidogenic activity, whereas ACTH(22-39) is less steroidogenic than ACTH(1-24)(16). If elevated ACTH levels have weak steroidogenic activity, ACTH(22-39) is the main elevated form. This matches the data showing that ACTH level elevation is dominant and cortisol level elevation is weak. The second possibility is the effluence of bioinactive ACTH from tumor tissue. Recent studies have indicated that paraneoplastic syndrome could be the pathophysiology of ICI-RH(17). Tumor tissues express ACTH or POMC ectopically as shared epitopes, and immunological responses toward not only the tumor tissue but also the pituitary gland cause ICI-RH(18). ICI administration may have damaged tumor cells, and tumor-derived ACTH may have been measured where it leaked into the bloodstream. Leaked ACTH/POMC, as a tumor-derived antigen, may activate antitumor activity, thereby causing ICI-RH as paraneoplastic autoimmune hypopituitarism. The third possibility is mild primary adrenal insufficiency before the onset of ICI-RH; subsequently, ICI-RH occurs as secondary adrenal insufficiency. When primary adrenal insufficiency such as adrenalitis occurs, ACTH concentration increases. The frequency of ICI-related primary adrenal insufficiency is lower than that of ICI-RH; however, this phenomenon occurs(19). If shared antigens between the normal adrenal gland and the pituitary exist, the immunological response toward the antigens may damage the adrenal gland first, which could be followed by damage of pituitary cells, thereby eliciting a transient elevation and a subsequent decrease in ACTH levels. Further prospective studies based on large case series are needed to clarify these mechanisms. Transient elevation of ACTH levels may provide insights into the pathogenesis of ICI-RH. In practice, we often need to evaluate ACTH and cortisol levels while modifying the history of glucocorticoid use. Monitoring fluctuations in ACTH levels could also be a marker of ICI-RH onset. However, ACTH fluctuations in patients with a history of steroid use are also an issue for future studies.

In conclusion, a transient elevation in ACTH levels occurred in some patients with ICI-RH before the onset of secondary adrenal insufficiency. Monitoring trends in ACTH levels could help predict the onset of ICI-RH.

## List of abbreviations

Ab: Antibody
ACTH: Adrenocorticotropic hormone
CTLA-4: Cytotoxic T-lymphocyte-associated protein 4
GNAL: Guanine nucleotide-binding protein G(olf) subunit alpha
HP: Hypothalamic pituitary adrenal
ICI: Immune checkpoint inhibitor
ICI-RH: ICI-related hypophysitis (ICI-RH)
irAE: immune-related adverse event
ITM2B: Integral membrane protein 2 B
PD-1, POMC: proopiomelanocortin, Programmed cell death protein 1
TSH: Thyroid-stimulating hormone

## Data Availability

All data produced in the present study are available upon reasonable request to the authors

## Declarations

### Ethics approval and consent to participate and publication

This study was approved by the ethics committee of the Kobe University Graduate School of Medicine (Approval No. 1685).

### Availability of data and material

Data are available on reasonable request.

### Authors’ contributions

All authors contributed to the article and approved the submitted version.

HB and GI conceived of and designed the study. HB and MY collected the data from electronic medical records. MY, YS, YOh, MK, YT, Yoi, and MT interpreted the data and contributed to discussion. HB performed the experiments and analyzed the data. HB wrote the manuscript and prepared all figures and tables. MY, SU, MS, NY, and GI edited the manuscript. HF and WO advised on the study and contributed to the critical revision of the manuscript for important intellectual content.

## Acknowledgements

We thank Y. Shindo, M. Nishime, H. Satoura, and K. Imura (Diabetes and Endocrinology) for their data collection and devotional patient care, and E. Maeda and Y. Kagawa (Medical Informatics and Bioinformatics) for their support in the extraction of data from electronic medical records. We would like to thank Editage (www.editage.jp) for English language editing.

